# Neurologic outcomes in people with multiple sclerosis treated with immune checkpoint inhibitors for oncologic indications

**DOI:** 10.1101/2024.06.13.24308608

**Authors:** Carson M. Quinn, Prashanth Rajarajan, Alexander Gill, Hannah Kopinsky, Andrew B. Wolf, Celeste Soares De Camargo, Jessica Lamb, Tamar Bacon, Joseph Murray, John C. Probasco, Kristin Galetta, Daniel Kantor, Patricia K. Coyle, Vikram Bhise, Enrique Alvarez, Sarah Conway, Shamik Bhattacharyya, Ilya Kister

## Abstract

**Objective:** To assess the prevalence of multiple sclerosis (MS) activity, as well as neurologic and non-neurologic immune-related adverse events (irAEs) in persons with MS (pwMS) treated with immune checkpoint inhibitors (ICIs) for cancer.

**Background:** ICIs are associated with irAEs and exacerbation of certain preexisting autoimmune diseases. PwMS are generally excluded from ICI clinical trials, so data on the safety of these drugs in pwMS is limited.

**Design/Methods:** Participating sites were recruited through the Medical Partnership 4 MS+ (MP4MS+) listserv. Seven large academic centers participated in the study, each conducting a systematic search of their electronic medical record system for patients with MS and ICI exposure. Demographics and data on MS and cancer history, treatments, and outcomes were abstracted from patient charts using a structured instrument.

**Results:** We identified 66 pwMS (median age 66 years, 73% female, 68% not on disease-modifying therapy (DMT) for MS) who were treated with ICIs for lung cancers (35%), melanoma (21%) or other oncologic indications. During post-ICI follow-up (median: 11.7 months, range 0.2-106.3 months), two pwMS (3%) had relapse or MRI activity, three (5%) had neurologic irAEs, and 21 (32%) had non-neurologic irAEs. At the last follow-up, 25 (38%) subjects had partial or complete remission of their cancer while 35 (53%) were deceased.

**Conclusions:** In this multi-institutional systematic retrospective study of predominantly older pwMS who were off DMTs, MS activity and neurologic irAEs following ICI treatment were rare, suggesting that preexisting MS should not preclude the use of ICIs.

## Introduction

Immune checkpoint inhibitors (ICIs) are increasingly used for a variety of cancers [1]. ICIs enhance the host anti-tumor immune response by blocking “immune checkpoints” – T-cell expressed programmed cell death protein 1 (PD-1), PD-1 ligand (PD-L1), and cytotoxic T lymphocyte antigen 4 (CTLA-4). A well-recognized side effect of immune upregulation is the development of immune-related adverse events (irAEs). It is estimated that 5% of irAEs involve the nervous system – neurologic irAEs, or nirAEs [2, 3] – manifesting as meningitis, encephalitis, myasthenia gravis (MG), myositis, Guillain–Barre Syndrome (GBS), and chronic peripheral neuropathies.

Patients with systemic preexisting autoimmune diseases have increased rates of irAEs and may experience worsening of their autoimmune disease [4]. Because typical MS syndromes, such as optic neuritis, transverse myelitis, and tumefactive demyelinating lesions, have been reported as nirAEs in patients on ICI, there is concern that the risk of these events would be increased in patients with preexisting MS [3, 5]. For this reason, pwMS were excluded from the major ICI trials and the safety profile of ICI use in pwMS is not well-characterized [6].

Initial case reports of pwMS receiving ICIs raised concerns about the risk of disease activity. Of the first 10 published cases of pwMS receiving ICIs for cancer treatment, 60% reported MS relapses after an average of 13.6 weeks, with symptoms of weakness, ataxia, optic neuritis, and brainstem syndromes [7]. However, only 2 of these 6 patients had poor neurologic outcomes, and in these 2 patients with poor outcomes, it was unclear if neurologic worsening was due to MS or other factors such as CNS radiation or nirAEs. Moreover, it is well recognized that case reports are biased toward worse outcomes, leading to overestimates of the risk of adverse events [6]. More recently, a few retrospective case series that assess the risk of relapse in MS following ICI have been published. In a multicenter case series from France, 18 pwMS who received ICIs were identified by treating clinicians. Clinical or radiographic worsening of MS was recorded in 3 patients (17%) within 6 months of ICI initiation. The authors noted that relapses occurred in younger patients who had stopped high-efficacy anti-trafficking DMTs, which are associated with disease rebound on discontinuation [8] and, therefore, may not be related to ICI. In a single-center study of 7 pwMS, no clinical relapses were reported following ICI start, but there were 2 cases of asymptomatic MRI lesions [9]. A case series from Brigham #x0026; Women’s Hospital also reported a very low rate of MS events [10]; the patients from this series were included in the current study. Reassuringly, although with limited data, pwMS also did not appear to have increased rates of irAEs, in contrast to what has been reported for other systemic autoimmune diseases [7, 11].

The aim of this study was to assess the rate of MS activity as well as of neurologic and non-neurologic irAEs in all ICI-treated pwMS from seven large academic centers across the United States.

## Methods

Participating sites were identified through the Medical Partnership 4 Multiple Sclerosis+ (MP4MS+) network, a platform for advocacy, clinical, and non-industry-funded research collaborations comprised of over 1,300 neurologists who treat pwMS. Investigators from the following institutions obtained IRB approval from their respective institutions and participated in the study: Mass General Brigham (Boston, MA), University of Colorado School of Medicine (Aurora, CO), Johns Hopkins Medicine (Baltimore, MD), Stanford University Medical Center (Palo Alto, CA), New York University Grossman School of Medicine (New York, NY), Stony Brook Medical Center (Stony Brook, NY), and Robert Wood Johnson Medical – Rutgers (Newark, NJ). Investigators at each site identified patients through institution-wide searches of the respective electronic medical record (EMR) system for patients with diagnosis codes of MS and treatment with any FDA-approved ICI (pembrolizumab, nivolumab, atezolizumab, cemiplimab, duravalamab, avelumab). We included all patients with a preexisting diagnosis of MS (confirmed by a neuroimmunologist at each site) who received at least one dose of an FDA-approved ICI medication for the treatment of cancer. Patients with Neuromyelitis Optica Spectrum Disease (NMOSD), Myelin Oligodendrocyte Glycoprotein Antibody Disease (MOGAD), Radiographically Isolated Syndrome, Clinically Isolated Syndrome, and those who developed MS after ICI start were excluded. Demographic and relevant clinical, laboratory, and radiologic data were extracted for each patient from the EMR using a comprehensive, structured instrument (Appendix 1). The key outcomes assessed were MS disease activity, nirAEs, non-neurologic irAEs following ICI initiation, and cancer outcomes. “MS disease activity” was defined as clinical relapse or new MRI lesions that could not be better attributed to CNS metastases or an alternative nirAE. MS activity was considered “related to ICI treatment” if it occurred within one year of the last ICI dose.

## Results

We identified 66 patients (73% female) with preexisting MS diagnosis who received at least one dose of ICI. The number of patients contributed by each site is shown in Figure 1. The median age at ICI initiation was 66.2 years (range 28.7-80.0 years). Most patients had inactive MS: 80.4% had no MS activity for ≥5 years, and 68.2% were off DMT at the time of ICI start. Details on the demographic and disease-related characteristics are shown in Table 1, and patients’ treatment histories are summarized in Table 2. Most patients received a single ICI, but 23% were on combination therapy. Importantly, 77% of patients were on cytotoxic chemotherapy at the time of ICI, and 21% had received cerebral radiation. The median duration of ICI treatment was 6.4 months (range: 1-39 months), with 17% of patients receiving only one cycle of ICI. The median post-ICI follow-up was 11.7 months. As expected, the follow-up was much shorter for deceased patients (n=35, median follow-up: 4 months) compared to those who were alive at the last follow-up (n=31, median follow-up: 24 months). The follow-up for each patient is shown in Figure 2.

**Table 1.** Baseline Characteristics.

|  | Count (%) / median (range) |
| --- | --- |
| Female | 48 (72.7%) |
| Age at ICI start (yrs) | 66.2 (28.7-80.0) |
| MS disease duration (yrs) | 20.8 (3.2-57.4) |
| Time between MS activity* & ICI start (yrs) | 9.0 (1.0-36.1) |
| DMT at ICI Start |  |
| No DMT | 45 (68.2%) |
| Glatiramer acetate | 8 (12.3%) |
| Interferon beta-1a | 5 (7.7%) |
| Dimethyl Fumarate | 3 (4.6%) |
| Rituximab | 2 (3.1%) |
| Fingolimod | 1 (1.5%) |
| Natalizumab | 1 (1.5%) |
| Ocrelizumab | 1 (1.5%) |
| Time b/w last DMT-ICI start (yrs) | 3.62 (0.2-20.4) |
| Functional Status at ICI Start |  |
| Ambulatory without support | 40 (60.6%) |
| Ambulatory with support (assist, cane, walker) | 19 (28.8%) |
| Bed or wheelchair bound | 7 (10.6%) |
| Primary Malignancy |  |
| Lung | 23 (34.8%) |
| Melanoma | 14 (21.2%) |
| Squamous Cell | 5 (7.6%) |
| Renal Cell | 4 (6.1%) |
| Urothelial | 4 (6.1%) |
| Breast | 3 (4.5%) |
| Other | 13 (19.7%) |
| CNS involvement of cancer at ICI start | 14 (22.6%) |

\*MRI lesion or clinical relapse

**Table 2.**
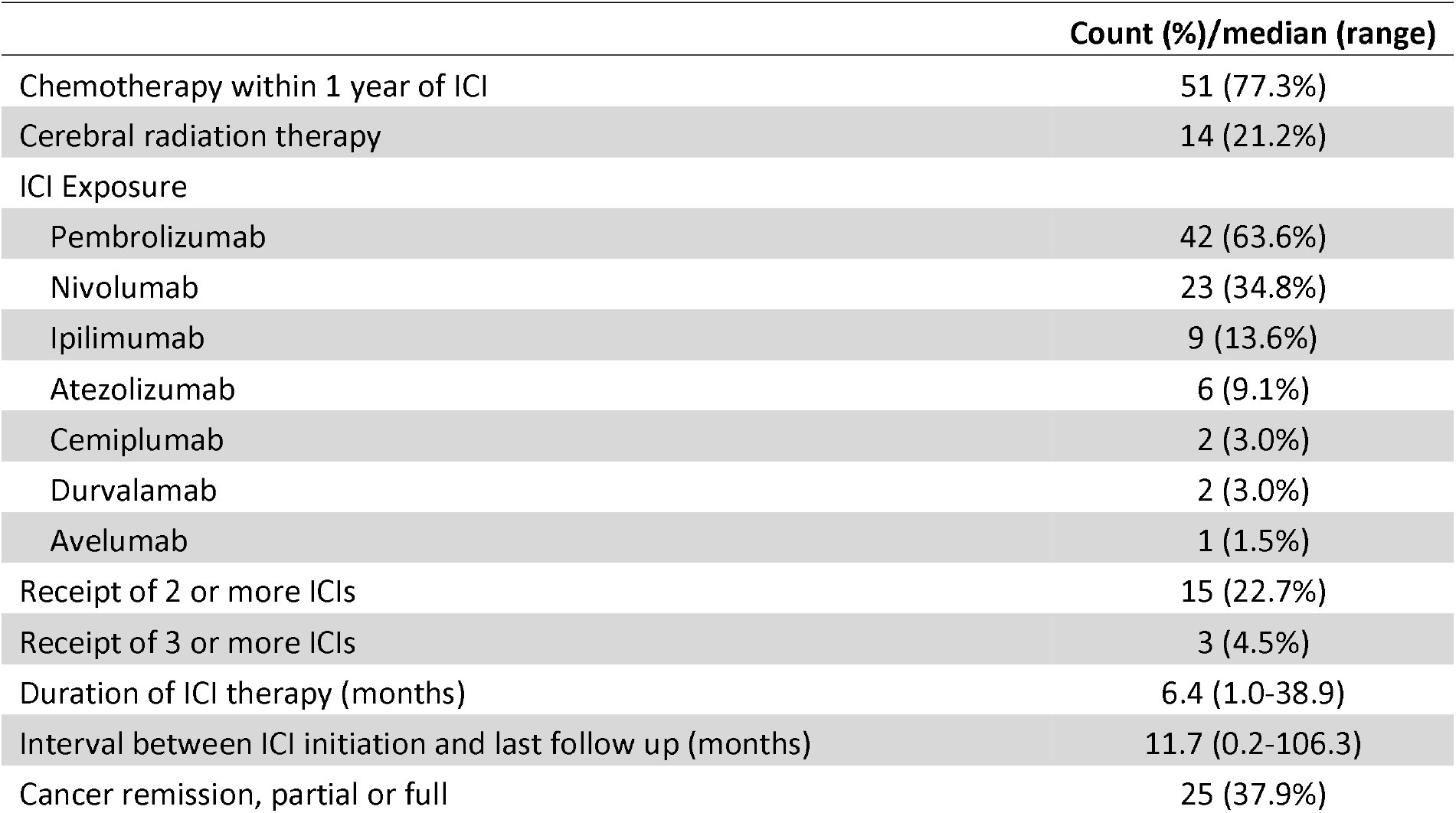
Cohort Cancer Treatment and Oncologic Outcomes.

|  | Count (%)/median (range) |
| --- | --- |
| Chemotherapy within 1 year of ICI | 51 (77.3%) |
| Cerebral radiation therapy | 14 (21.2%) |
| ICI Exposure |  |
| Pembrolizumab | 42 (63.6%) |
| Nivolumab | 23 (34.8%) |
| Ipilimumab | 9 (13.6%) |
| Atezolizumab | 6 (9.1%) |
| Cemiplumab | 2 (3.0%) |
| Durvalamab | 2 (3.0%) |
| Avelumab | 1 (1.5%) |
| Receipt of 2 or more ICIs | 15 (22.7%) |
| Receipt of 3 or more ICIs | 3 (4.5%) |
| Duration of ICI therapy (months) | 6.4 (1.0-38.9) |
| Interval between ICI initiation and last follow up (months) | 11.7 (0.2-106.3) |
| Cancer remission, partial or full | 25 (37.9%) |

**Table 3.**
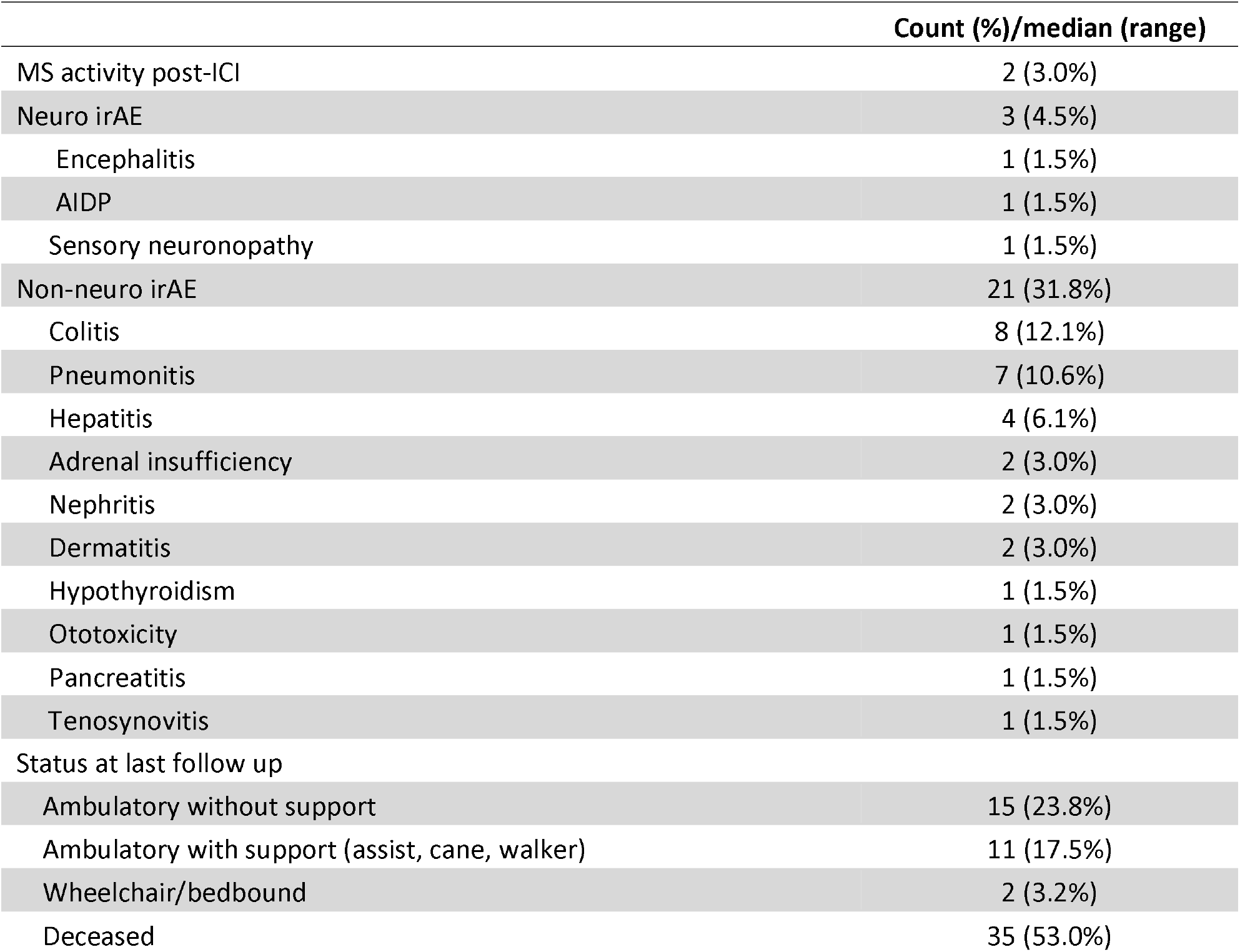
Outcomes – adverse events.

**Figure 1.**
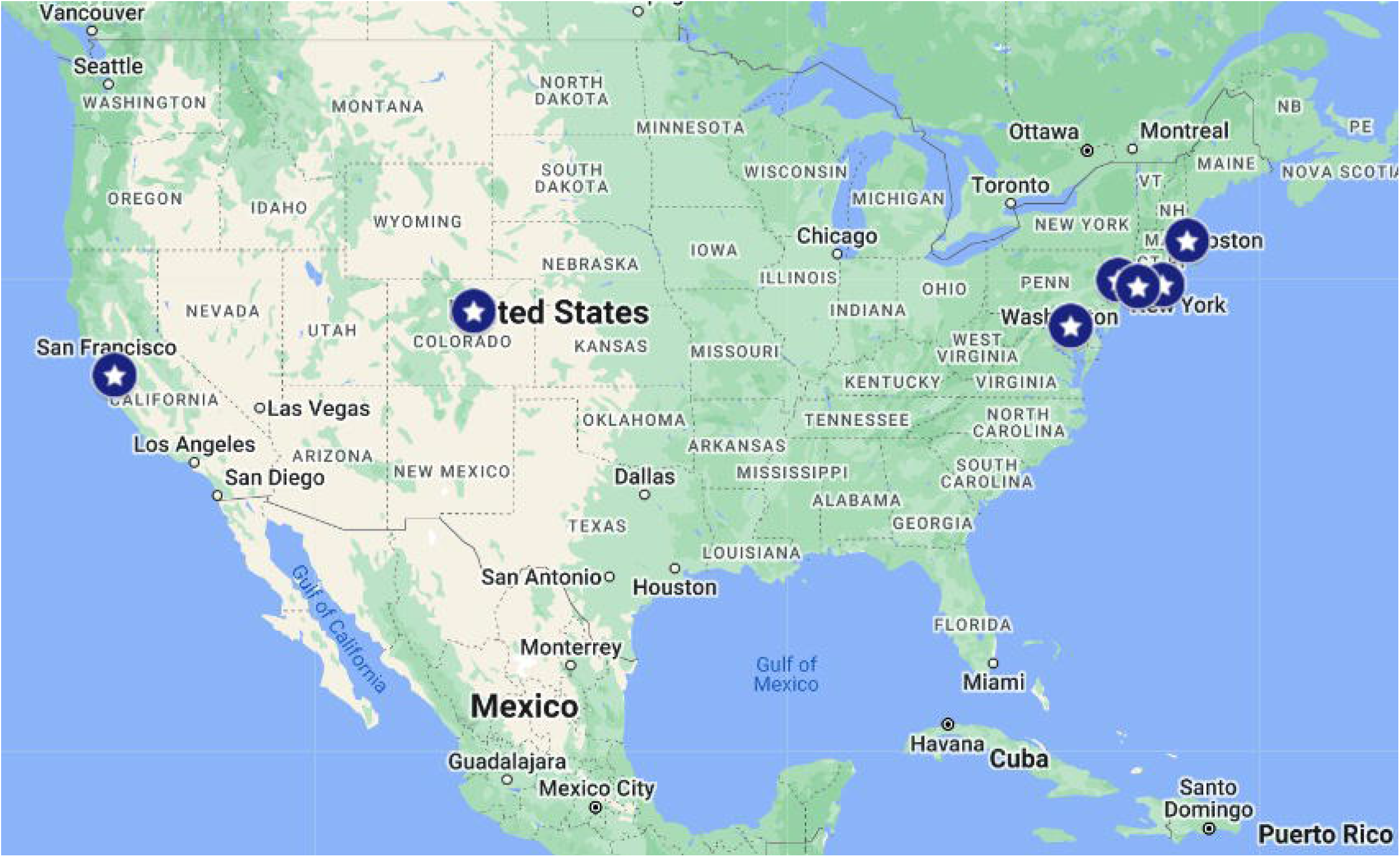
Map of sites in the MP4MS+ network which contributed patients to the cohort.

**Figure 2.**
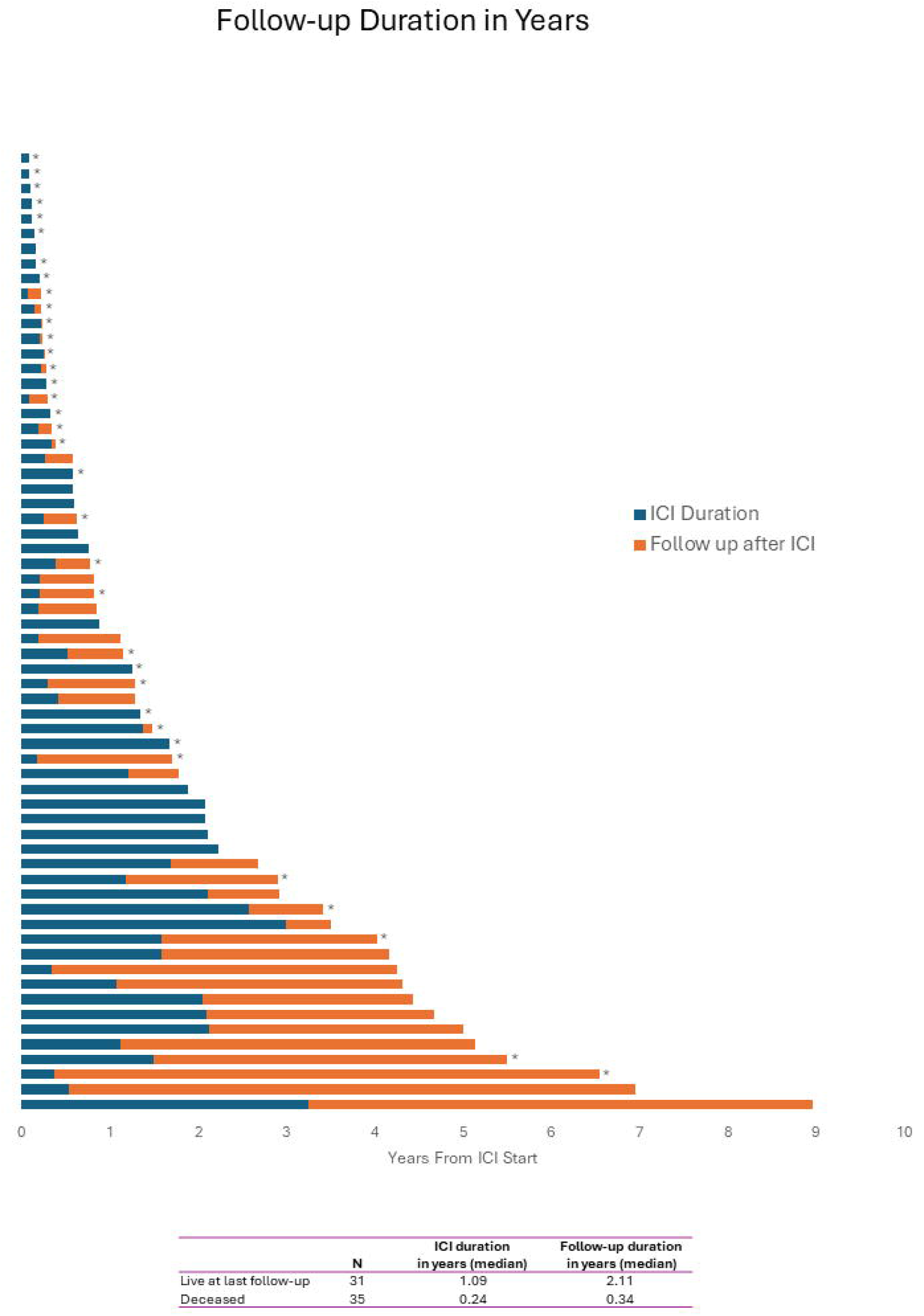
ICI-treatment duration and follow-up. A swimmer plot depicting ICI-treatment duration (blue) and subsequent follow-up time (orange) in years for each patient. Patients that were deceased at the time of last follow-up are marked with an asterisk (*). In total, of the 31 patients alive at last follow up, there was a median follow up of 2.11 years; and of the 35 deceased at last follow up, there was a median follow up duration of 0.34 years.

MS disease activity within 1 year of ICI start was recorded for only 2 patients (3%). One male patient aged 60-65 years, without MS activity for greater than five years at the time of ICI start experienced a mild clinical relapse with enhancing MRI lesions and worsening gait that resolved with corticosteroids. The second patient (female, aged 60-65 years), without MS activity for greater than five years, who had two small asymptomatic T2 hyperintense lesions on the MRI brain 4 months after ICI start. Both were on DMT at the time of the new activity. One additional patient had radiographic MS disease activity, which was deemed unrelated to ICI as it occurred 15 months after cessation of ICI therapy. Detailed timelines for the three cases with MS activity are shown in Figure 3a.

**Figure 3.**
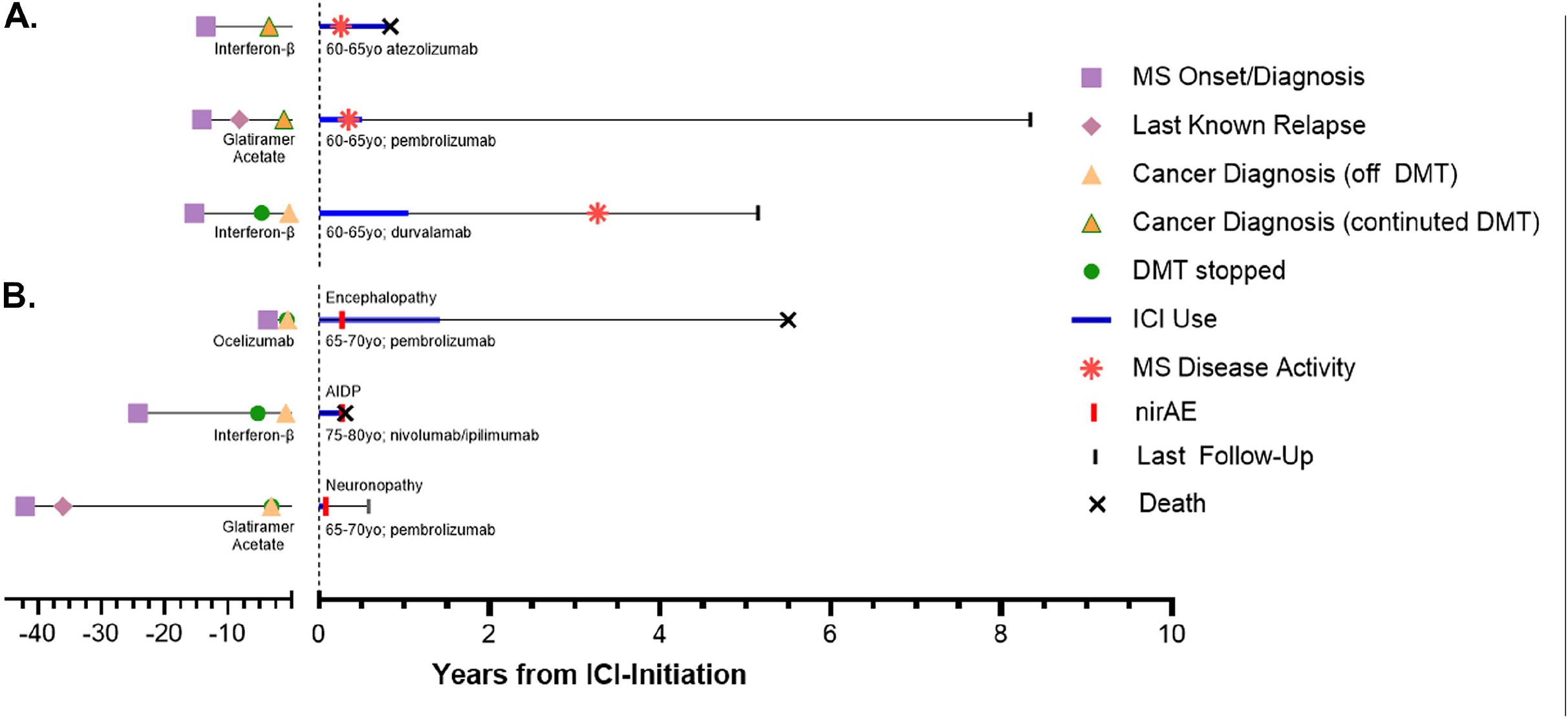
MS disease activity and neurologic irAEs clinical timeline. Timeline in years of patients with (A) MS disease activity and (B) neurologic irAEs within one year ICI-treatment. Of note, a third patient with MS disease activity post-ICI treatment is depicted, although this activity was more than one year after cessation of ICI-therapy.

As younger patients tend to be at higher risk of MS relapses, we conducted a sub-analysis of patients in our series who were 50 years or younger at the time of ICI start. We identified 6 such patients, but, despite younger age, they also tended to have inactive MS, with a median of 5.3 years (range 2.3-8.3 years) between the last MS activity and ICI start. None of the 6 patients was on DMT at ICI start, though 3 had only recently discontinued treatment with interferon beta-1a, rituximab, and ocrelizumab. None of the 6 experienced MS activity or nirAEs during follow-up.

### Three patients had nirAEs

one had encephalitis with seizures, one had a fatal case of Guillain-Barré Syndrome (despite treatment with high-dose intravenous corticosteroids and immunoglobulin [IVIG]), and one developed sensory neuronopathy. The timelines for these three patients are shown in Figure 3b. Non-neurologic irAEs were reported for 21 patients (32%) and were most commonly colitis, pneumonitis, and hepatitis (Figure 4).

**Figure 4.**
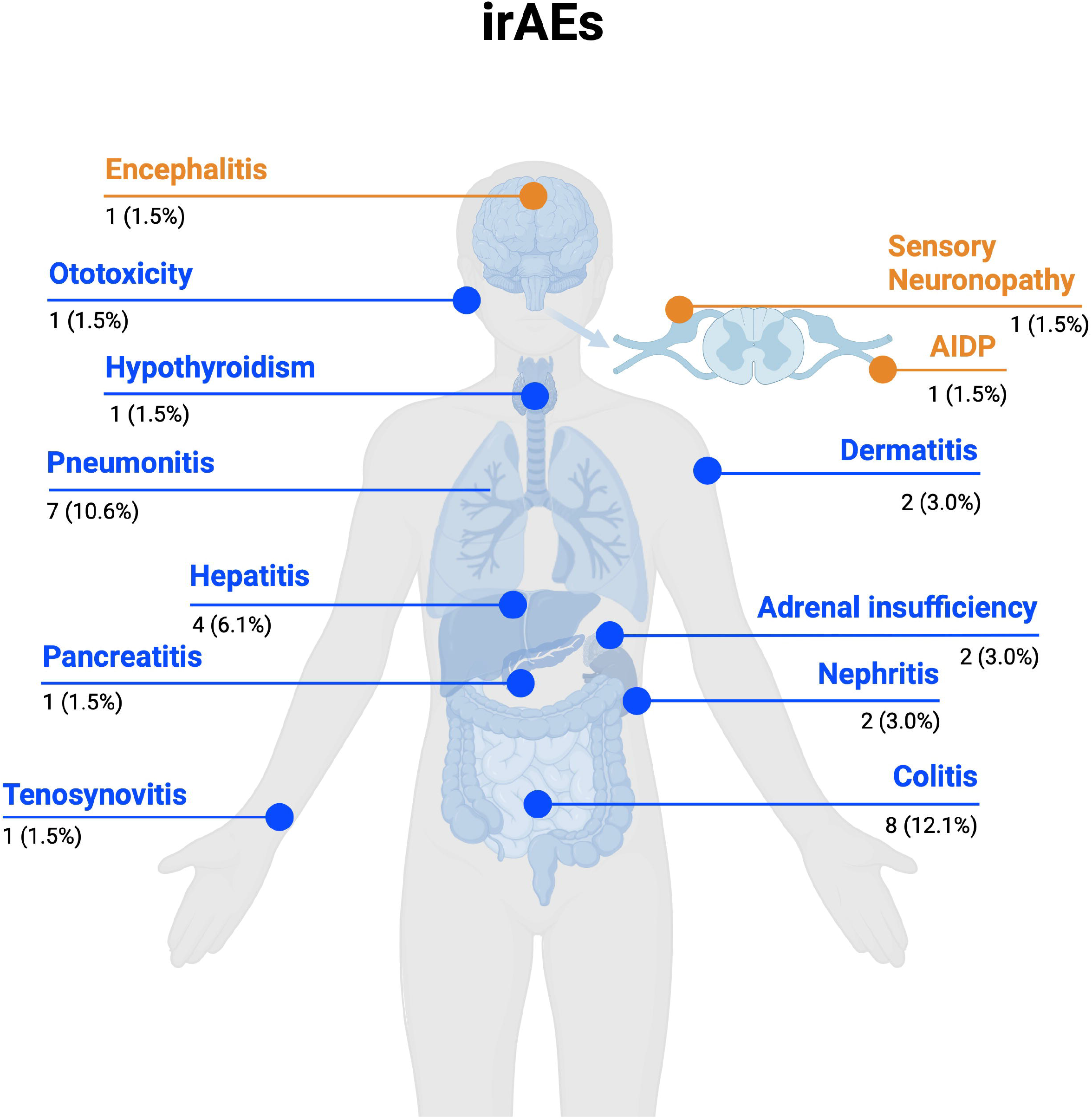
Immune-related adverse events (irAEs) in people with MS. Schematic depicting all observed neurologic (orange) and non-neurologic (blue) irAEs observed in this cohort of people with MS exposed to ICI-therapy. Numbers beneath irAE type depict the number (percentage) of patients with each ICI-associated condition.

As of the last follow-up, 38% of patients had complete or partial cancer remission, 53% were deceased, and the remainder either did not achieve remission or were missing data. Of the patients who were alive at the last follow-up, 48% were ambulatory without assistance.

## Discussion

In our multicenter case series of 66 pwMS receiving ICIs for cancer, MS activity was observed in 3% of patients. This rate is similar to what would be expected in inactive MS patients of similar age who are not on ICI therapy [12]. We did not observe any patient with “disease rebound” (disease activity out of proportion to what has been observed prior to ICI). On the contrary, both MS events in our series were mild – one patient had asymptomatic MRI lesions, and the other had gait impairment responsive to corticosteroid treatment. In a recently published series from France, MS activity rate was numerically higher – in 3 out of 18 patients (17%) – than in our series; however, their patients were younger (median 48 vs 66 years in our series) and MS activity in two of the three patients in the French series occurred in the setting of discontinuation of natalizumab, a high-efficacy MS DMT associated with MS rebound [7, 8, 13]. Thus, post-ICI relapses in the French series could be related to stopping anti-trafficking DMT rather than to ICI. In our series, only 2 patients stopped anti-trafficking agents (fingolimod and natalizumab), but neither experienced any MS activity. A smaller, single-center series from UCSF reported no relapses in 7 patients [9]. Taken together, the three case series that examined MS activity after ICI therapy, including ours, comprise a total of 91 pwMS, of whom 2 (2.2%) had clinical relapses, and 5 (5.5%) had asymptomatic MRI activity [8, 9].

Importantly, we examined not only MS activity in our patients but also rates of neurologic and non-neurologic irAEs. We found that the rate of nirAEs (5%) was similar to non-MS patients receiving ICI in the clinical trials, consistent with prior series [3]. The rate of non-neurologic irAE was 31.8% in our patients, which is within the range of what is expected in non-MS patients treated with ICIs [4]. Cancer remission was achieved in a minority of our patients (38%), while the majority were deceased at the last follow-up (53%), reflective of the advanced stage of their malignancies at the time of ICI initiation. Whether preexisting MS had delayed time to ICI initiation and whether earlier ICI use could have improved outcomes cannot be determined with our data but warrants further investigation.

Why some autoimmune diseases predispose patients to higher-than-expected rates of preexisting disease exacerbation while MS does not is unknown [4]. Perhaps a higher degree of immune dysregulation in systemic autoimmune disorders compared to MS predisposes the former to ICI-related immune complications as well as to other immune diseases [14]. However, this is unlikely to be the only reason, as patients with other purely neurologic autoimmune diseases, such as MG, appear to be at high risk for ICI-related relapse [15]. At present, despite considerable efforts to identify biomarkers that would predict irAEs in patients who receive ICI, no biomarker has been validated for clinical use [16]. Further work to better understand the immunopathogenesis of irAEs may shed light on why certain conditions show a greater predisposition to this complication and lead to the development of reliable biomarkers of treatment-emergent autoimmunity that would make ICIs safer.

Our study has several strengths. The model of using the broad network of MP4MS+ allowed us to identify multiple participating academic institutions across the US, resulting in the largest series of MS patients exposed to ICI – more than doubling the total number of cases of ICI in pwMS published to date. All investigators conducted comprehensive EMR searches that included all eligible patients from their institution, which reduces the risk of bias, whereby the more severe outcomes are more likely to be recalled and reported. Our median post-ICI follow-up was approximately a year, and 51 (78%) of patients had follow-up for 3 months, which is the period when most irAEs are reported.

Our study shares the well-known limitations of retrospective design, including missing data, loss to follow-up, and lack of controls [17]. Patients receiving ICI typically have advanced cancer and high morbidity despite aggressive therapy. Thus, patients may not be able to follow up with their treating neurologist and receive a neurologic examination. Lacking such data, we are not able to comment on whether ICI has an impact on disease progression independent of relapse activity (PIRA). This question will need to be addressed in future studies, ideally with prospective follow-up. It is also important to note that not all patients had brain MRIs after ICI initiation – 39% had missing MRI data – and it is possible that asymptomatic lesions may have been missed. However, minor asymptomatic MRI changes do not typically have long-term implications for disability [18]. Lastly, because a minority of patients were deceased within 3 months of onset, which is the window for the emergence of most of the treatment-related irAEs, it is not possible to know what their MS course would have been like had they lived longer.

It is important to emphasize that our patients were older relative to the typical MS clinic population, mostly had inactive MS for many years, and were off DMTs. Our conclusions about the lack of MS activity post-ICI may not be generalizable to younger pwMS with more active MS or those who stop anti-trafficking DMT (anti-integrin monoclonal antibodies, S1P receptor modulators) that are associated with disease rebound on discontinuation. Larger multicenter cohorts, inclusive of younger MS patients with more active disease, are needed to confirm and extend our findings. Moreover, most patients in our cohort were on prior or concomitant cytotoxic chemotherapy, which may reduce the risk of relapse and potentially offset the tendency to ICI-associated relapses. We are not able to comment on whether ICI would be similarly safe – from an MS standpoint – in pwMS who start ICI prior to receiving cytotoxic chemotherapy. Lastly, it should be emphasized that conclusions about the apparent safety of ICI in MS cannot be extrapolated to other neuroinflammatory conditions with different pathogenesis, such as NMOSD or MOGAD. Efforts are currently underway to explore the safety of ICI in these rare neuroinflammatory disorders, with preliminary evidence pointing to a different risk profile with ICI [19].

Despite the caveats, our data is reassuring in that older patients with inactive MS could receive ICI treatment for cancer without the risk of MS reactivation. Our findings argue that older pwMS should not be excluded from future clinical trials of ICIs. In younger patients, who may be at higher risk for relapses, it may be prudent to continue highly effective MS therapy while on ICI if this does not interfere with oncologic treatment. Special care is necessary to weigh the pros and cons of stopping anti-trafficking therapy prior to ICI, which may lead to disease rebound (independent of ICI treatment). Ideally, pwMS who initiate ICI should be under the care of a multidisciplinary team that includes a neurologist experienced with MS.

## Data Availability

Deidentified data produced in the present study are available upon reasonable request to the authors

